# Estimating Physician-Patient Face-to-Face Time Using Electronic Health Record Audit Logs

**DOI:** 10.64898/2026.10.10.26364993

**Authors:** Nate C. Apathy, Dori A. Cross

**Affiliations:** Department of Health Policy and Management, School of Public Health, University of Maryland, College Park, Maryland; Division of Health Policy and Management, School of Public Health, University of Minnesota, Minneapolis, Minnesota

**Keywords:** electronic health records, ambulatory care, EHR audit logs, measure agreement, physician fee schedule, face-to-face time

## Abstract

**Importance:** The Medicare Physician Fee Schedule values ambulatory services using survey-based estimates of physician time, a costly process vulnerable to inaccuracy. EHR audit logs are passively and universally collected and likely meet the American Medical Association Relative Value Scale Update Committee (AMA RUC) Extant Database Standards for alternative data to inform service valuation estimates. However, audit logs’ validity for estimating face-to-face time, a core component of many of the most used procedural codes, is unknown.

**Objective:** To assess whether EHR audit logs can facilitate accurate estimation of physician-patient face-to-face time during ambulatory visits.

**Design:** Single-site validation study pairing EHR audit logs with direct audio/video recordings of ambulatory primary care visits (reference standard). We assessed agreement between audit-log estimates and recorded visit duration with Bland-Altman analysis and mean absolute error.

**Setting:** University of Pennsylvania Observer data repository, a multi-modal database of ambulatory visits to Penn Medicine primary care clinics including video and audio recordings, transcripts, clinical EHR data, and EHR audit-log data.

**Participants:** 75 visits from 12 physicians with data collected between August 2023 and February 2025.

**Exposures:** A rule-based algorithm estimated face-to-face time via clusters of physician EHR activity relative to patient check-in time. Inactivity thresholds were applied to differentiate intraservice time with no EHR activity (e.g., a physical exam) from the end of a visit, with longer thresholds used for longer visits.

**Main Outcomes and Measures:** Duration of recorded face-to-face visit time.

**Results:** Recorded face-to-face time averaged 31.6 minutes (SD, 13.9). The audit-log estimator was unbiased (mean bias: -0.63 minutes; 95% CI: -6.0 to 2.6; mean absolute error: 9.0 minutes) and estimated face-to-face time within approximately 2% (38 seconds) of the recorded visit time with no proportional bias (slope: 0.13; P = .25). The 95% limits of agreement were -25.7 to 24.4 minutes, and 37% of estimates fell within 10% of recorded duration.

**Conclusions and Relevance:** In this validation study, EHR audit logs recovered unbiased estimates of ambulatory face-to-face time. Larger-scale studies are required to verify generalizability, but EHR audit logs offer a scalable means to improve the rigor of retrospective physician work relative value unit estimation. This study represents a first step in a potential paradigm shift for measuring components of the fee schedule across procedures, settings, and provider types.

**Key Points:** *Question:* Can electronic health record (EHR) audit log data be used to accurately estimate physician-patient face-to-face time for ambulatory visits?

*Findings:* In this single-site validation study of 75 ambulatory visits with paired visit recordings, a combined rule-based estimation strategy recovered average face-to-face time to within approximately 2% of recorded duration (mean bias: -38 seconds), with no significant proportional bias, although visit-level agreement was modest.

*Meaning:* Universally and passively collected EHR audit log data may provide a scalable, low-bias source for estimating discrete components of physician service time to inform fee-schedule valuation.

---

Accurately valuing physician services for payment is critical to a high-performing health care system. In the US, the Medicare Physician Fee Schedule (PFS) allocates billions of dollars annually to pay physicians based on Work Relative Value Units (wRVUs), which are derived primarily from survey-based physician estimates of service time and complexity and subsequently assigned to Current Procedural Terminology (CPT) codes.^1^ This costly, labor-intensive process is vulnerable to mis-estimation and may contribute to systemic inaccuracies in how physician services are valued.^2,3^ As a result, the Centers for Medicare and Medicaid Services (CMS) has expressed a desire to improve the accuracy of service time estimation deployed for wRVU components specifically (e.g., estimating physician pre- and intra-service time) and other components of CPT code valuation (e.g., estimating practice expense inputs).^2,4–6^ However, given the strict standards in place for extant, observational data used in CPT code valuation, few data sets can readily qualify.^1^ Thus far, much of the progress in examining alternative data for CPT valuation has focused on surgical settings because of the availability of anesthesia claims and surgical logs to capture operative intra-service time.^7–13^ Expanding this work into the most commonly billed ambulatory Evaluation and Management (E/M) CPT codes has the potential to impact a far larger volume of CMS spending across more specialties, but data sources have proven elusive. However, data from electronic health record (EHR) audit logs, passively collected click-level records capturing all activity in a patient’s chart, are likely to contain both the granularity and broad availability necessary to improve the accuracy of service time estimation for ambulatory care.^14–17^

Ambulatory E/M visits are heavily brokered through clinician use of the EHR,^18,19^ making EHR audit log data a particularly useful resource for proxying how time is spent during these types of visits. Audit logs are also a mandatory function for all EHR vendor systems certified by the Office of the National Coordinator for Health IT (ONC), primarily for use in compliance activity (e.g., auditing inappropriate chart access). These data are universally and passively collected and therefore not subject to manual manipulation or data entry errors, in addition to being both attributable to individual EHR users and linkable to individual patient visits. Two studies have successfully used EHR-derived activity timestamps (but not formal audit logs) to estimate visit duration and intraservice time in ambulatory settings, finding discrepancies between their measures and current benchmarks.^20,21^ However, neither study had the ability to validate estimates, which is crucial for understanding the accuracy and potential biases of any novel estimation approach. Establishing the reliability and credibility of audit log-derived methods in a baseline encounter context with ground truth validation can help to build the analytic foundation to support analysis of large, representative visit samples and variation across specialties, patient complexity, and visit types. These important potential sources of variation are not currently reflected in the PFS and could inform policies to ameliorate under-investment in cognitive specialties relative to procedural specialties.^3,22,23^

The aim of this study is to validate the accuracy and reliability of using EHR audit log data^24^ to estimate face-to-face time for ambulatory visits. We focus on face-to-face time because it is the largest wRVU time component for the most frequently used CPT codes in ambulatory care. We do this by comparing encounter-specific time estimates from a simple audit log-derived face-to-face time estimator to the duration of paired video recordings for our sample of visits. This study offers important insight into potentially scalable, validated solutions for addressing a long-standing challenge of recovering accurate intraservice time estimates from extant, universally collected data to inform payment rates for ambulatory services.

## Methods

### Data Source: Penn Observer Repository

This study followed the Guidelines for Reporting Reliability and Agreement Studies (GRRAS).^25^ The study was deemed non-human subjects research by the University of Maryland – College Park IRB due to the use of deidentified data. EHR audit log data were paired with deidentified visit recordings from the University of Pennsylvania Observer Repository, a novel multimodal dataset of ambulatory primary care visits.^24^ For each visit, the repository includes (1) EHR audit log data capturing time-stamped user actions in the record and (2) an audio/video recording of the entirety of the patient-physician clinic visit interaction. This recording duration provides a direct, time-and-motion style observed estimate of physician-patient face-to-face time. Visit recordings are not derived from an ambient listening tool that physicians may turn on or off throughout their time with the patient. Patients and clinicians provided consent to have the entire encounter video- and audio-recorded. More information about the methods used to compile the Observer repository are available in prior published work.^24^

### EHR Audit Log Data

The core data for the estimation algorithm is derived from the EHR audit log, a data source that is increasingly used in health services research and health informatics given its rich representation of detailed user activity.^14,16,17,26^ The audit log captures (roughly) click-level activity for all users in the EHR, meaning that actions taken by any user that touches a patient’s chart (viewing a record, modifying, etc.) are recorded with a precise timestamp. The Observer audit log data includes all audit log actions by all users, including visit physicians, for a given patient visit.

### Physician-Patient Face-to-Face Time Estimation Algorithm

We estimated each visit’s face-to-face time based on physician activity in each patient’s chart. We restricted the audit log data for each visit to that of only the visit physician, and excluded all activity prior to patient check-in, which is recorded via a registration workflow and appears as a discrete logged event for every visit (metric 15137 in Epic’s ACCESS_LOG). This ensured that any pre-charting that occurred before the patient’s arrival in the clinic was not included in our estimates.

Physician EHR use within an encounter frequently demonstrates bursts of EHR activity separated by brief spans of inactivity.^27^ For each visit’s set of physician audit log actions, we first identified and grouped clusters of EHR activity, calculating the interstitial time between each action and each cluster. In our estimates of cumulative face-to-face time, we wanted shorter spans of EHR inactivity to be considered inclusively, as these windows frequently represent direct interactions with the patient like a physical exam. Beyond a certain window of elapsed time, however, the absence of EHR activity is more likely to signal the end of an encounter rather than an intra-visit pause. This time period is commonly referred to as an “inactivity threshold” in EHR audit log research.^17,27^ Based on the observation from visit recordings that longer patient visits tend to involve more time-consuming exams and discussions with patients and family members that may be present, we opted to calibrate our estimation algorithm, and appropriate inactivity threshold, separately for shorter versus longer visits.

With discrete EHR activity clusters identified and segmented via inactivity thresholds (described in further detail below), face-to-face time estimates were computed as the elapsed time span between the first and last action in the longest-duration session of activity without surpassing the inactivity threshold for the focal visit. An annotated visual depiction of the estimation algorithm for two example visits including annotations of inactivity thresholds is provided in **Figure 1**.

**Figure 1.**
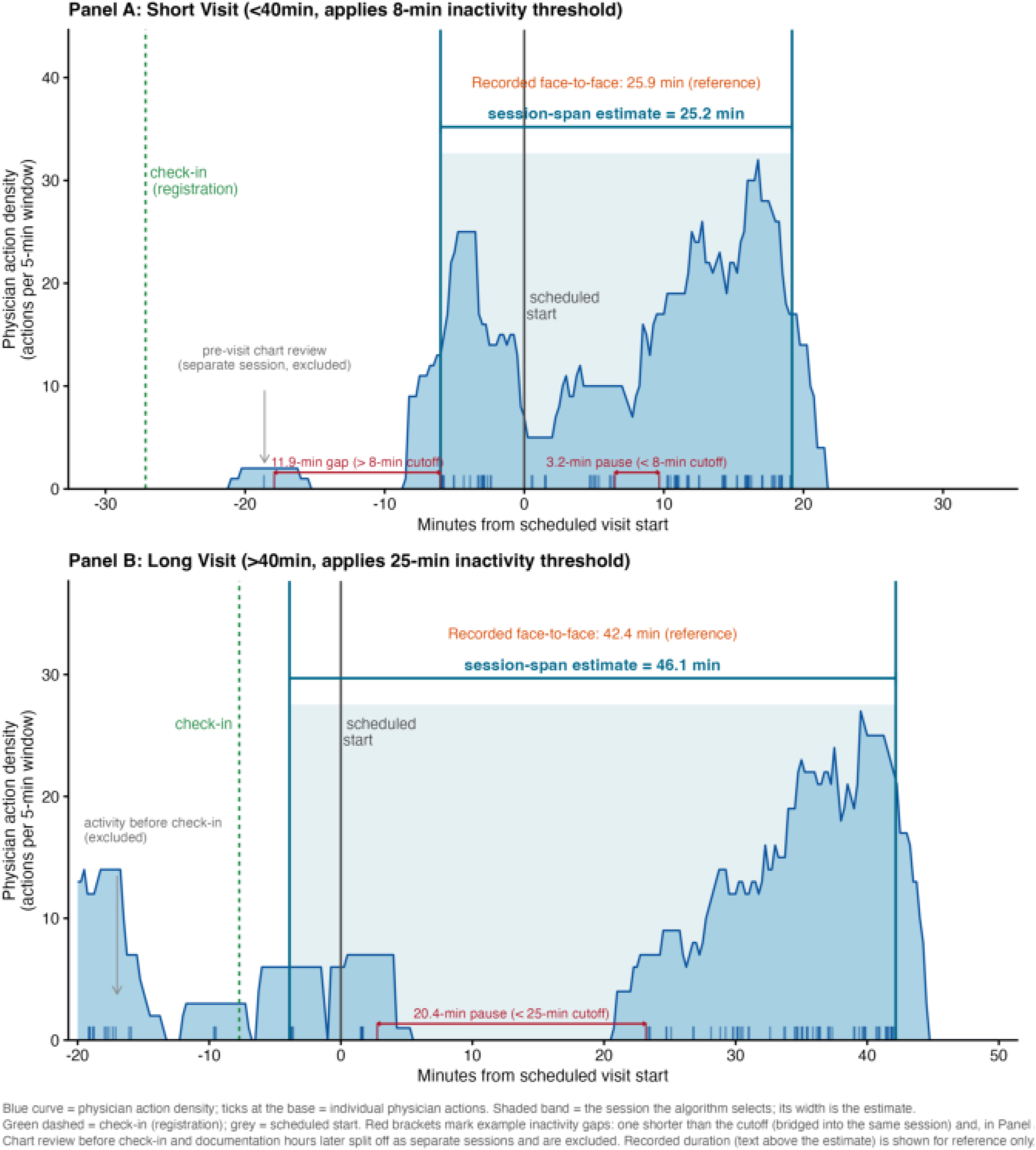
Illustration of the audit-log face-to-face time estimation algorithm for two example visits. Each panel plots the visit physician s EHR action density (actions per 5-minute window) against time relative to the scheduled visit start (0) and patient check-in (registration). The shaded band is the session the algorithm selects —the longest post check-in span in which no inactivity gap exceeds the length-conditional threshold —and its width is the estimate. Red brackets mark example inactivity gaps: one below the threshold (bridged into the session) and, in Panel A, one above it (splitting pre-visit chart review into a separate, excluded session). A, short visit (<40 minutes; 8-minute threshold); B, long visit (≥40 minutes; 25-minute threshold).

Importantly, our estimation algorithm does not require physicians to be actively using the EHR for time to “count” towards the estimated face-to-face time (note the gaps between physician EHR activity in **Figure 1**). The algorithm does, however, rely on the assumption that any interaction that the physician has face-to-face with the patient is *bookended* or delimited by logged EHR activity. Any logged activity by the physician in the patient’s chart, including chart closure, qualifies for this bookending as long as this terminal EHR activity falls within the inactivity threshold.

### Statistical Analysis

To assess agreement between audit-log derived estimates and the recorded visit duration, we used Bland-Altman (B-A) analysis,^28–30^ the standard for assessing agreement between two methods of clinical measurement. Because both measures carry error, measuring correlation alone is insufficient, as the two measures can be strongly associated but disagree systematically.^30^ Consistent with B-A, the reported measures include the average bias (defined as the mean difference between the audit-log estimate and the recording duration), the physician-clustered 95% confidence interval of the bias via bootstrap resampling of physicians (5,000 replications), and the 95% limits of agreement. Each visit’s difference (estimate minus recording duration) was plotted against the mean of the audit-log-derived estimate and recording duration. We tested for proportional bias by regressing the difference on this mean. This analysis was complemented with two additional measures of performance: error magnitude (via mean absolute error as well as root mean squared error), and the proportion of estimates within a pre-specified tolerance (±10% of the visit recording duration).

### Inactivity Threshold Determination

Inactivity thresholds were set differently for short and long visits because extended encounters had longer intra-service windows of non-EHR use within the face-to-face visit period (i.e., a lengthier conversation). We used data-driven cut-points based on recorded visit durations to define short visits as those lasting less than 40 minutes (the 80^th^ percentile of recording duration) and long visits as 40+ minutes. We then tested inactivity thresholds ranging from 2 minutes to 40 minutes in both short and long visit subsamples to identify the inactivity threshold(s) that minimized the estimation algorithm’s mean bias for each subsample of visits. Mean bias was minimized with an 8-minute inactivity threshold for short visits and a 25-minute inactivity threshold for long visits, consistent with our expectations of needing distinct inactivity thresholds for visits of varying scheduled durations (**eFigure1**). All analyses were performed in R, version 4.5.2 (R Project for Statistical Computing).

## Results

The analytic sample comprised 75 visits with paired recordings and EHR audit log data contributed by 12 physicians (median of 5 encounters per physician, maximum of 20). Recorded face-to-face time averaged 31.6 minutes (SD, 13.9; **Table 1**).

**Table 1.** Distribution of face-to-face time: audit log estimate vs recording reference.

| Measure | Mean (min) | SD | Median | Min | Max |
| --- | --- | --- | --- | --- | --- |
| Recording duration (reference) | 31.6 | 13.9 | 28.7 | 9 | 70.4 |
| Audit log estimate | 31.0 | 15.5 | 27.9 | 3.1 | 83.6 |

SD, standard deviation.

### Face-to-face time estimation

Our algorithm was unbiased in estimating the recorded face-to-face time, with a mean bias of -0.63 minutes (95% CI: -6.0 to 2.6) and a mean absolute error (MAE) of 9.0 minutes (**Table 2**). Large underestimates were more common than large over-estimates (**Figure 2**). In B-A analysis, the 95% limit of agreement was -25.7 to 24.4 minutes with no significant proportional bias (slope: 0.13; 95% CI: -0.09 to 0.35; P =□.25; **Figure 3**). 37.3% of audit-log estimates were within 10% of the recorded duration, highlighting that despite relatively low performance in per-visit prediction, aggregate estimates can have low bias.

**Table 2.**
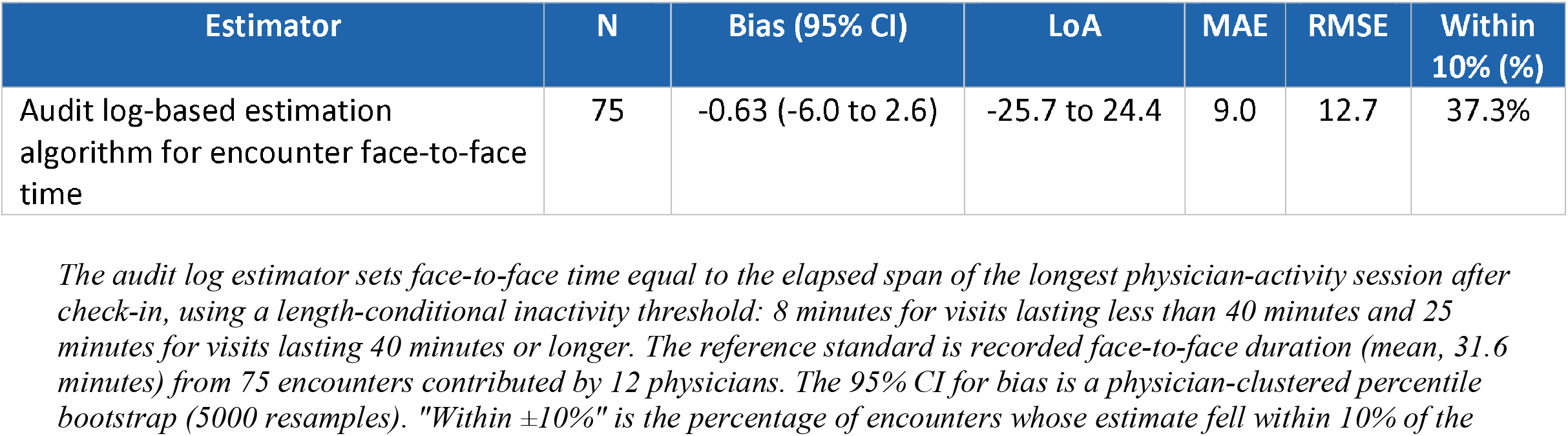
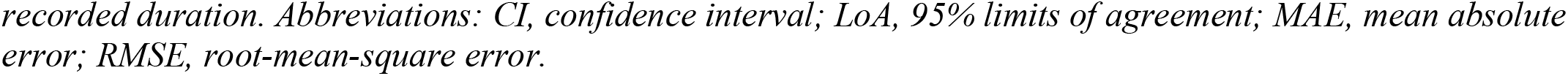
Agreement between audit log estimates and recorded face-to-face time.

| Estimator | N | Bias (95% CI) | LoA | MAE | RMSE | Within 10% (%) |
| --- | --- | --- | --- | --- | --- | --- |
| Audit log-based estimation algorithm for encounter face-to-face time | 75 | -0.63 (-6.0 to 2.6) | -25.7 to 24.4 | 9.0 | 12.7 | 37.3% |

*The audit log estimator sets face-to-face time equal to the elapsed span of the longest physician-activity session after check-in, using a length-conditional inactivity threshold: 8 minutes for visits lasting less than 40 minutes and 25 minutes for visits lasting 40 minutes or longer. The reference standard is recorded face-to-face duration (mean, 31.6 minutes) from 75 encounters contributed by 12 physicians. The 95% CI for bias is a physician-clustered percentile bootstrap (5000 resamples). "Within $\pm 10\%$ " is the percentage of encounters whose estimate fell within 10% of the*
recorded duration. Abbreviations: CI, confidence interval; LoA, 95% limits of agreement; MAE, mean absolute error; RMSE, root-mean-square error.

**Figure 2.**
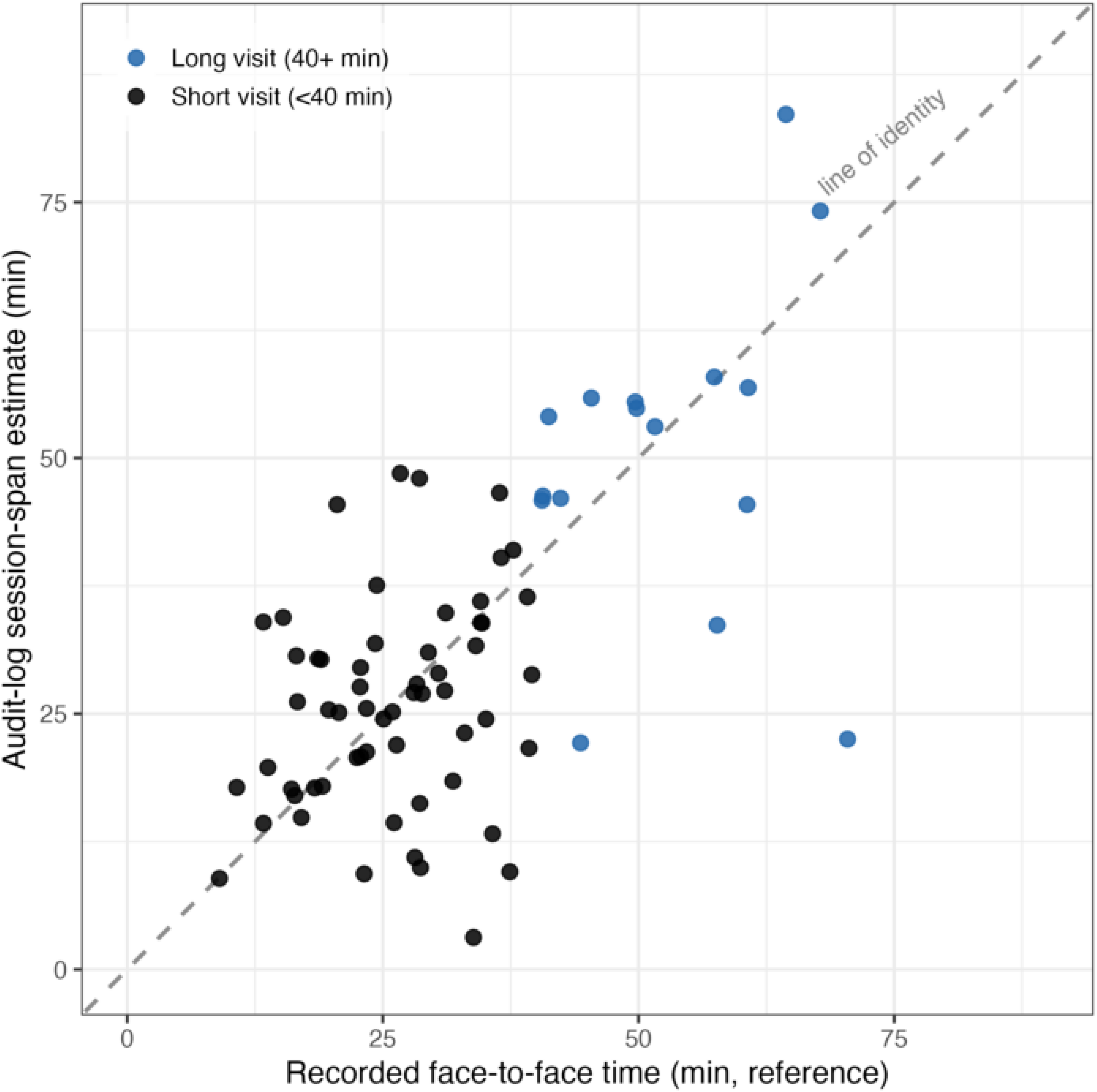
Audit-log estimated versus recorded face-to-face time for 75 ambulatory visits. Each point is one visit; the diagonal is the line of identity (estimate = recording). Points are colored by stratum (short, <40 minutes; long, ≥40 minutes).

**Figure 3.**
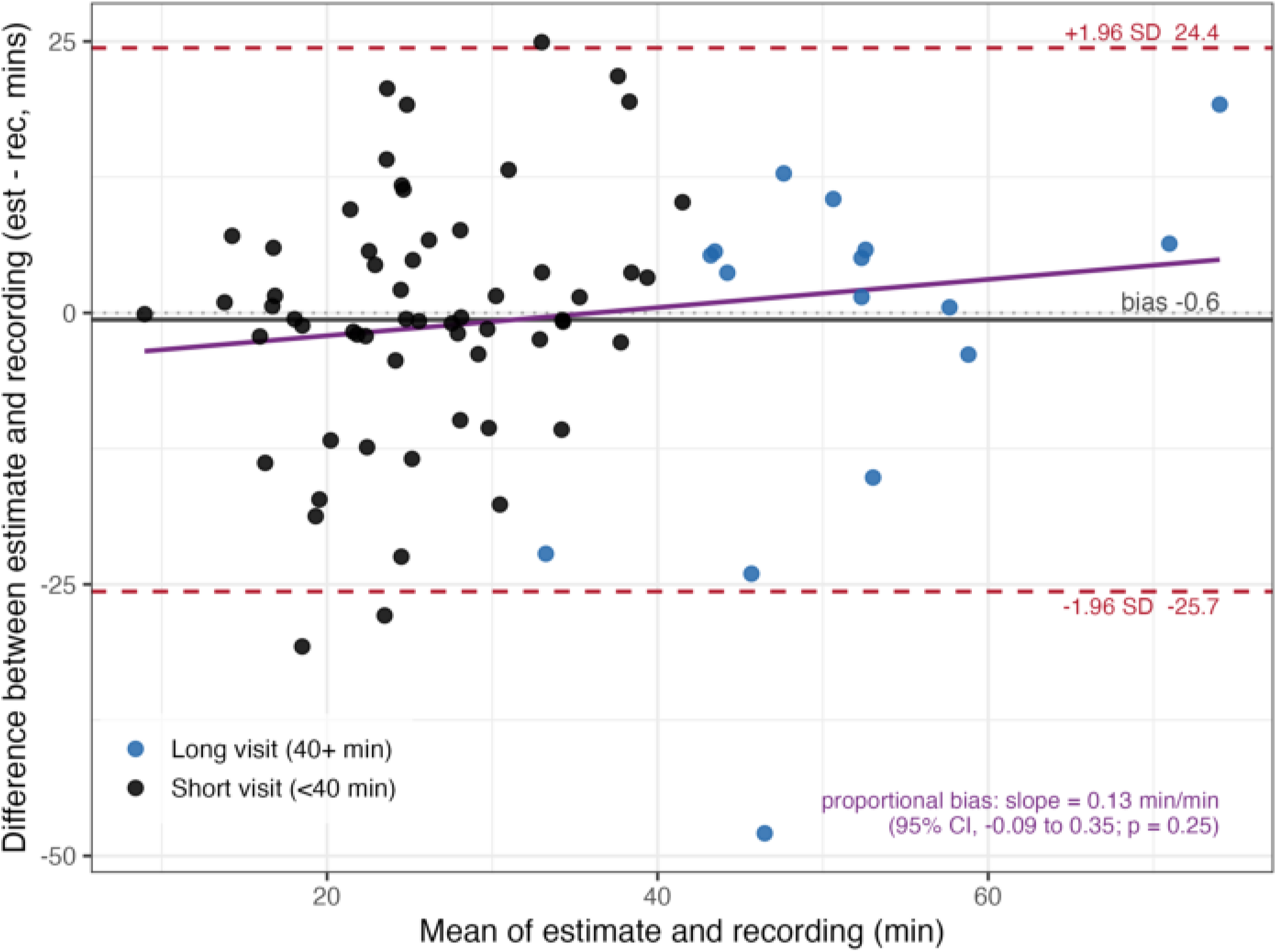
Bland-Altman plot of agreement between audit-log estimates and recorded face-to-face time (N = 75). The difference (estimate − recording) is plotted against the mean of the two measures for each visit. The solid line is the mean bias (−0.6 minutes); dashed lines are the 95% limits of agreement (−25.7 to 24.4 minutes); the diagonal line is the proportional-bias regression of the difference on the mean (slope, 0.13 min/min; 95% CI, −0.09 to 0.35; P = .25).

## Discussion

This study demonstrates the feasibility and validity of estimating physician-patient face-to-face time via EHR audit logs, recovering estimates that are on average within 38 seconds or 2.0% of the average face-to-face time captured in visit recordings. To date, this is the first validated demonstration that EHR audit log data can recover unbiased average estimates of face-to-face time for ambulatory visits, and the first to apply EHR audit log data specifically to the estimation of the discrete components of physician service-times that underpin wRVUs. The algorithmic approach to face-to-face time estimates developed here can be readily applied to other EHR audit log data sets, perhaps paired with audio recording durations from widely adopted ambient scribes. The analytical design decisions require no pre-existing knowledge or measurement of physician EHR audit log activity patterns. Additionally, the only conditional logic required to appropriately assess and apply different inactivity thresholds relies simply on being able to parse visit observations by short versus long duration. For additional validation work, this could continue to rely on recorded visit duration as source of truth; if these validated methods become widespread, future work could test the method’s performance using *scheduled* visit durations.

It is important to emphasize that the value of this method for policymaking lies in its ability to recover unbiased estimates of *average* face-to-face time across a sample of visits, rather than attempting to estimate the exact duration of any one visit. As illustrated by the wide limits of agreement (±25 minutes) and relatively low proportion of estimates within 10% of the recorded duration (37%), this method is not well-suited for measuring face-to-face time for individual visits. Rather, its policy relevance lies in the recovery of unbiased aggregate parameters that take advantage of random (and thus offsetting) errors for individual encounters. There is well-established precedent in health policy for similar approaches that demonstrate low accuracy for any single unit but perform well in aggregate. For example, CMS Hierarchical Condition Categories (HCC) risk-adjustment models are used to estimate predicted expenditures for large groups of beneficiaries in Medicare Advantage plans based on patient demographics and diagnostic criteria.^31^ While these models have poor unit-level predictive performance (explaining only about 10% of the beneficiary-level variation in expenditures), these models are considered acceptable for continued use based on their high predictive ratio, which is the ratio of average predicted expenditures to actual average expenditures for the entire group of beneficiaries.^31,32^ For purposes of comparison, the predictive ratio for our method of estimating face-to-face time is 0.981 (31.0 minutes / 31.6 minutes, with 1.0 being perfect prediction). As a result, this method is promising for the policy question we aim to address: informing CPT code valuation using population estimates of service time for the typical instance of a given CPT code across a large sample of visits.^1^

This study extends a small but growing literature aiming to improve the rigor applied to estimation of physician service time for ambulatory visits via widely collected secondary data.^20,21^ In the ambulatory space, estimation of physicians’ time spent actively working in the EHR has been studied extensively with aggregated EHR use metadata capturing users’ active use time (e.g., Epic Signal or Oracle Health’s Advance Platform).^16,18,33–39^ While EHR use metadata has also been used to extrapolate physician work time estimates,^40,41^ it consists of aggregated data designed with a fundamentally different purpose of characterizing EHR-based activity rather than estimating totality of visit duration. It also does not currently meet the requirements of extant data to inform the PFS in that it is not linkable to specific visits or, in turn, CPT codes.^1^ Therefore, our study represents three important innovations: first, we demonstrate the methodological feasibility of deriving low-bias estimates of face-to-face time in ambulatory care; second, we validate these estimates against an observed duration of the face-to-face time; and finally, our method uses broadly available, universally collected data (EHR audit logs) that are likely to meet the criteria put forth by the RUC for extant data for use in RBRVS recommendations.^1,4^

There are several natural extensions to this work that future research can explore. First, replication studies are required to ensure that this method is generalizable across additional users, specialties, and organizations. Larger-scale studies may leverage recordings from widely used ambient listening tools or artificial intelligence (AI) scribes^42–44^ for a reference value, although quality checks will be required to ensure that these recordings capture the entirety of physician-patient face-to-face time. Second, RVUs are comprised of a host of *time-based estimates* in addition to intraservice or face-to-face time. Pre-service time, post-service time, staff time spent on tasks like collecting vital signs data, and many other temporal constructs are ripe for rigorous empirical examination at scale supported by EHR audit logs. Moreover, the EHR audit log should be explored further to investigate methods to modernize other *non-time-based* dimensions of physician fee-setting and prices (e.g., by assessment of visit volume patterns during post-surgical global billing periods). While data alone cannot supplant expert clinical judgment, estimates from EHR audit logs may serve as empirical anchors for rate-setting discussions.

### Limitations

This study has several limitations. First, all data in this study comes from a single, large, academically affiliated health system, which may limit the generalizability of the results. Second, while observed visit recording duration is used to bifurcate the sample and apply different inactivity thresholds for face-to-face time estimation, the recording duration may not necessarily directly correspond to the scheduled visit duration for these visits (this data is not currently available in the Observer repository for this sample). As a result, agreement using scheduled rather than observed visit durations requires separate validation. Third, the sample is relatively small for a validation study, so our study may be leveraged primarily as establishment of the novel face-to-face time estimation method for future, larger-scale studies. Fourth, with only one recording per visit, reference-standard error cannot be definitively distinguished from estimator error, although visit recordings in this sample are very likely to be high-fidelity representations of face-to-face visit time. Finally, the audit logs in the Observer repository are *visit-centric*, and thus already parsed in a way that an organization’s raw audit log data is not. Initial processing of the raw file (containing all logs for all patients by all clinicians) will require some algorithmic modification and may exhibit better (or worse) agreement. For example, sessions that are delimited by changes in the patient context (i.e., the same user closing patient A’s chart and opening patient B’s) are not observable in the Observer audit logs but could easily be constructed and tested in a future validation study.

## Conclusion

This validation study developed and applied a simple rule-based algorithm using EHR audit logs to reliably estimate the average physician-patient face-to-face time for ambulatory visits within 2.0% of the recorded duration. The findings illustrate the potential utility of EHR audit logs in supporting scalable, rigorous estimation of discrete components of physician service time. Further research is required to validate this approach across sites and specialties and to expand the application of these methods and data sources to additional components of the fee schedule. EHR audit log data may offer a scalable, universal, and prospectively collected alternative data source to improve the rigor of physician work RVU estimation.

## Supporting information

eFigure

## Data Availability

All data is available from the Penn Observer Repository upon completion of a data use agreement with the University of Pennsylvania.

https://observer.med.upenn.edu

## Data availability

All data is from the University of Pennsylvania Observer Repository (https://observer.med.upenn.edu/). Details about data access requests are available at that site. Code for estimation replication will be posted publicly on the author’s GitHub upon publication.

## Author contributions (CRediT)

**Nate C Apathy**: Conceptualization, Data curation, Formal analysis, Funding acquisition, Investigation, Methodology, Project administration, Validation, Visualization, Writing - Original Draft, Writing - Review & Editing; **Dori A Cross**: Methodology, Supervision, Writing - Review & Editing;

## Prior Presentation

An earlier version of this work was presented at the AMIA Clinical Informatics Conference, May 2026.

