## Supplementary material for "Estimating Physician-Patient Face-to-Face Time Using Electronic Health Record Audit Logs": eFigure

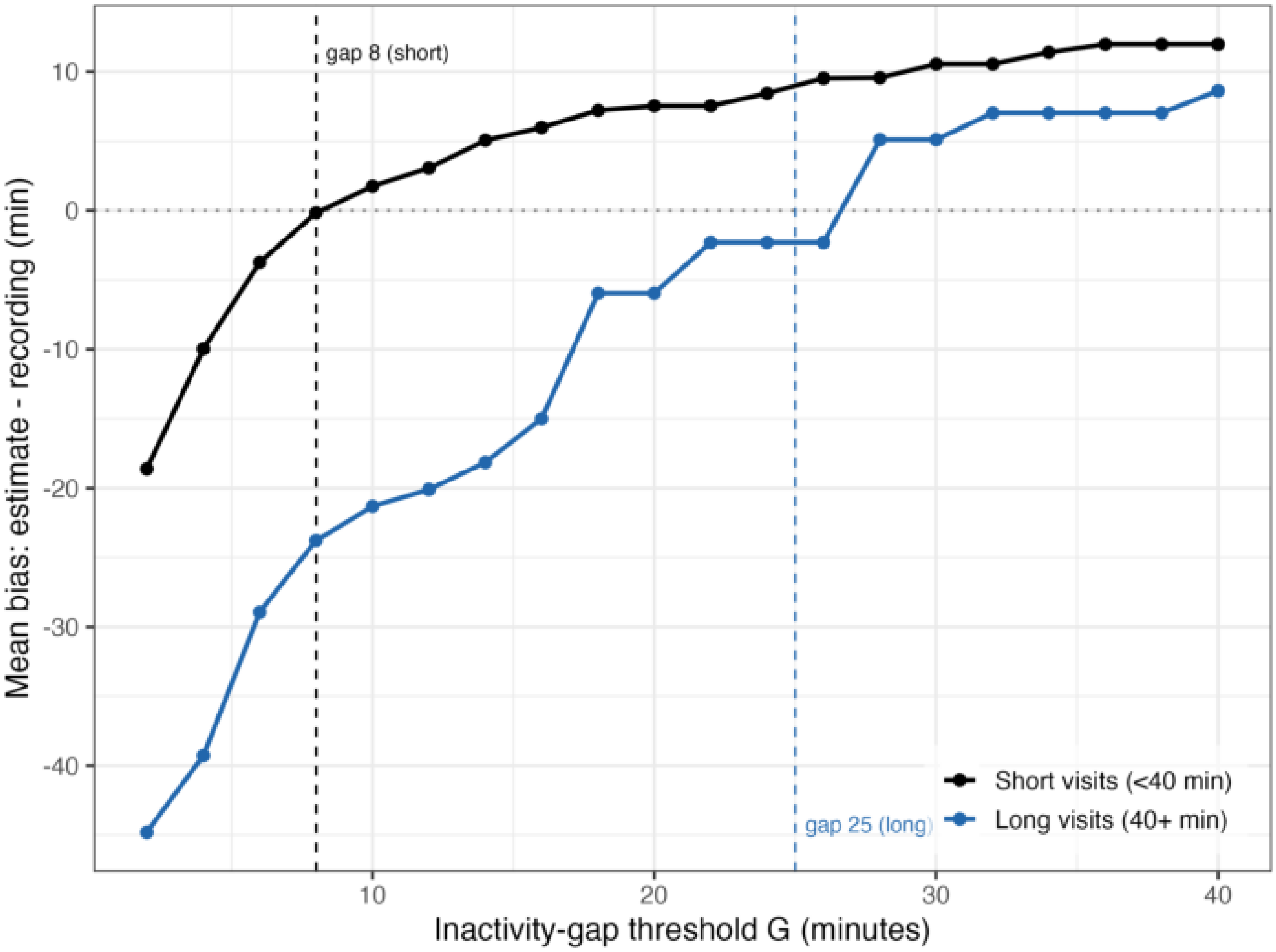


**eFigure 1.**
Inactivity-threshold tuning. Mean bias of the audit-log estimator (estimate − recorded duration, in minutes) as a function of the inactivity-gap threshold, computed separately for short visits (<40 minutes) and long visits (≥40 minutes). Thresholds were varied from 2 to 40 minutes; the values minimizing absolute mean bias were 8 minutes for short visits and 25 minutes for long visits—the thresholds used in the length-conditional primary analysis.
